# *MAPT* allele and haplotype frequencies in Nigerian Africans: population distribution and association with Parkinson’s disease risk and age at onset

**DOI:** 10.1101/2023.03.24.23287684

**Authors:** Olaitan Okunoye, Oluwadamilola Ojo, Oladunni Abiodun, Sani Abubakar, Charles Achoru, Olaleye Adeniji, Osigwe Agabi, Uchechi Agulanna, Rufus Akinyemi, Mohammed Ali, Ifeyinwa Ani-Osheku, Owotemu Arigbodi, Abiodun Bello, Cyril Erameh, Temitope Farombi, Michael Fawale, Frank Imarhiagbe, Emmanuel Iwuozo, Morenikeji Komolafe, Paul Nwani, Ernest Nwazor, Yakub Nyandaiti, Yahaya Obiabo, Olanike Odeniyi, Francis Odiase, Francis Ojini, Gerald Onwuegbuzie, Godwin Osaigbovo, Nosakhare Osemwegie, Olajumoke Oshinaike, Folajimi Otubogun, Shyngle Oyakhire, Simon Ozomma, Sarah Samuel, Funmilola Taiwo, Kolawole Wahab, Yusuf Zubair, Dena Hernandez, Sara Bandres-Ciga, Cornelis Blauwendraat, Andrew Singleton, Henry Houlden, John Hardy, Mie Rizig, Njideka Okubadejo

**Author notes:** **Correspondence** Njideka U. Okubadejo, Neurology Unit, Department of Medicine, Faculty of Clinical Sciences, College of Medicine, University of Lagos, Lagos State, Nigeria.

## Abstract

**Background:** The microtubule-associated protein tau (*MAPT*) gene is critical because of its putative role in the causal pathway of neurodegenerative diseases including Parkinson’s disease (PD). However, there is a lack of clarity regarding the link between the main H1 haplotype and risk of PD. Inconsistencies in reported association may be driven by genetic variability in the populations studied to date. Data on *MAPT* haplotype frequencies in the general population and association studies exploring the role of *MAPT* haplotypes in conferring PD risk in black Africans are lacking.

**Objectives:** To determine the frequencies of *MAPT* haplotypes and explore the role of the H1 haplotype as a risk factor for PD risk and age at onset in Nigerian Africans.

**Methods:** The haplotype and genotype frequencies of *MAPT* rs1052553 were analysed using PCR-based KASP™ in 907 individuals with PD and 1,022 age-matched neurologically normal controls from the Nigeria Parkinson’s Disease Research (NPDR) network cohort. Clinical data related to PD included age at study, age at onset, and disease duration.

**Results:** The frequency of the main *MAPT* H1 haplotype in this cohort was 98.7% in individuals with PD, and 99.1% in healthy controls (p=0.19). The H2 haplotype was present in 41/1929 (2.1%) of the cohort (PD - 1.3%; Controls - 0.9%; p=0.24). The most frequent *MAPT* genotype was H1H1 (PD - 97.5%, controls - 98.2%). The H1 haplotype was not associated with PD risk after accounting for gender and age at onset (Odds ratio for H1/H1 vs H1/H2 and H2/H2: 0.68 (95% CI:0.39-1.28); p=0.23).

**Conclusions:** Our findings support previous studies that report a low frequency of the *MAPT* H2 haplotype in black ancestry Africans, but document its occurrence in the Nigerian population (2.1%). In this cohort of black Africans with PD, the *MAPT* H1 haplotype was not associated with an increased risk or age at onset of PD.

## Introduction

The microtubule-associated protein tau (*MAPT*) gene located on chromosome 17q21 encodes the tau protein which is primarily involved in modulating the stability of axonal microtubules within the brain.^1^ *MAPT* has two distinct haplogroups (H1 and H2) resulting from inversion of an approximately 900kb region, and which, while having the same amino acid sequence, demonstrate polymorphic variability capable of altering gene expression (including transcription, post-transcriptional modification, and rates of translation).^2^ H1 (the non-inverted sequence) is predominant, whereas the less common H2 (the inverted haplotype) is reported most frequently in Europeans and Southwest Asians with a frequency of 23.5 – 37.5%.^3^ In general, the lowest rates in published literature are from the sub-Saharan African population (0.7% - 6.3%) and East Asians/Pacific Islanders (almost 0%).^3^ Notably, the proportion of African samples included in these reports have been low, typically below 50 participants within any African ethnic group, and subject to sampling error and underestimation.

Mutations in *MAPT* are associated with neurodegenerative tauopathies including Alzheimer’s disease (AD), progressive supranuclear palsy (PSP), and fronto-temporal dementia (FTD), but have also been implicated in Parkinson’s disease (PD) which is predominantly a synucleinopathy. ^4^ Whereas normal tau is integral to maintaining neuronal functioning, stabilizing and assembling microtubules, regulating axonal transport, promoting neurite growth, and preventing DNA damage and apoptosis, abnormal hyperphosphorylated tau (the product of *MAPT* mutations) promotes neurodegeneration.^4^ Conformational change, increased tendency to misfold, dissociation from mictrotubules, aggregation into neurofibrillary tangles, and ultimate loss of normal function characterize the main pathological process in tauopathies.^5, 6^ Clarifying the role of *MAPT* in neurodegenerative disorders has implications for developing neuroprotective interventions to reduce or inhibit tau phosphorylation, aggregation, and promote microtubule stabilization. The H1 haplotype has been postulated to contribute to an increased risk of PD, whereas H2, correlating with reduced tau protein expression, may be protective.^1, 7-11^ Studies of the association between *MAPT* and risk of PD have demonstrated different effect sizes (and suggested a weak effect), the possibility of interactions with other PD associated susceptibility loci, and ethnic variability even within populations of similar geographical origin. ^8-13^ A highly significant association between PD and the H1/H1 haplotype was documented in the Genetic Epidemiology of Parkinson’s Disease (GEO-PD) consortium study that included 5302 Caucasians with PD of Caucasian.^11^ The effect was independent of any interaction with *SNCA*, applied to risk of PD, but with no significant effect on the age at onset of PD.^11^

In the present study, the main objectives are to describe the population distribution of *MAPT* main haplotypes in Nigerian Africans, and thereby fill the void regarding the frequencies of the *MAPT* haplotypes in a large population of sub-Saharan Africans of black ancestry, and secondly, determine if there is an association between *MAPT* haplotype variation and the risk of, and age at onset of PD in our population.

## Methods

### Study design and participant recruitment

Study participants in this cohort study were recruited by participating neurologists in the Nigeria Parkinson’s Disease Research (NPDR) network as part of an ongoing cohort study conducted in collaboration with the International Parkinson’s Disease Genomic Consortium-Africa (IPDGC-Africa).^14,15^ The NPDR cohort is nationally representative and includes participating sites in all 6 geopolitical zones of Nigeria. Research ethics approval for the study was obtained from the the institutional health research ethics committees, the National Health Research Ethics Committee (NHREC) in Nigeria and the University College London (UCL) Institutional Review Board. All participants provided written informed consent.

Individuals with PD fulfilled the United Kingdom Parkinson’s Disease Society Brain Bank (UKPDSBB) criteria (the only exception being that we did not exclude those with a first degree relative who also had PD).^16^ Controls were otherwise healthy volunteers with no known family history of PD and no clinically evident neurological condition, from the same population, and matched for age. Baseline demographic characteristics (age at study, gender, age at onset of PD, and duration of PD (years)) were documented.

### *MAPT* genotyping

DNA was extracted either from saliva samples collected using DNA Genotek® saliva kits or from venous whole blood samples using standard protocols. *MAPT* locus haplotypes were investigated by tagging the H1/H2 haplotypes with the major allele and minor allele of SNP rs1052553. For rs1052553 allele A and G correspond to H1 and H2 haplotypes, respectively.^17^ Genotyping was performed using the Kompetitive Allele-Specific Polymerase Chain Reaction (PCR) assay (KASP™, LGC Genomics. Herts, UK). Genotypes were assessed for Hardy-Weinberg equilibrium (HWE) using Fisher exact test.

### Statistical analysis

Cohort characteristics and *MAPT* rs1052553 haplotype and genotype frequencies were expressed as proportions (%) compared between groups (individuals with PD and controls) using two tailed ?2 test. Power calculations were performed using the GAS Power calculation tool (http://csg.sph.umich.edu/abecasis/cats/gas_power_calculator/) using the following parameters: disease prevalence of 1%, N cases = 910, N controls = 1022, allele frequency = 0.05, genotype relative risk = 1.5, and a significance level of 0.05. Our analyses showed an 80% statistical power to detect an OR of ∼1.48.

Logistic regression analysis was applied to investigate the association between *MAPT* haplotypes and PD risk and age of onset. For all analyses, PD (cases) and/or controls were the dependent variable, with the relevant *MAPT* haplotype or genotype as the independent variable(s), adjusting where relevant for sex, age at onset or at age at study in PD and age at recruitment for controls. For descriptive purposes, age at onset of PD and age at study for controls were categorized by 10-year intervals and disease duration by 5-year intervals. Data were analysed using Stata/MP version 16.0 statistical software (Stata Corporation, College Station, TX: StataCorp LLC).

## Results

### Baseline characteristics of study population

In this study, we included 907 Nigerians with PD and 1,022 neurologically healthy controls matched for age (p=0.10). Median duration of PD was 3.0 years. The baseline characteristics of the cohort (sex distribution, age at study, age at onset, duration of PD overall and by gender) are shown in Supplementary Table 1. Genotype frequencies were consistent with Hardy-Weinberg equilibrium (HWE) (p>0.78).

### *MAPT* allele/haplotype and genotype frequencies in PD compared to controls

The *MAPT* rs1052553 allele/haplotype and genotype frequencies are shown in Table 1 and did not differ significantly between individuals with PD and controls (either overall or by gender). The most frequent allele was the major allele A (H1) with a haplotype frequency of 98.7% in PD and 99.1% controls (p=0.19). The most frequent genotype was H1/H1 in both PD (884, 97.5%) and controls (1004, 98.2%) (p=0.24). *MAPT* H2 (with number of participants as denominator) was present in 2.1% (41/1929) of the entire cohort, 2.5% (23/907) in PD and 1.76% (18/1022) in controls (see Table 1 footnote). Within the Nigerian ethnic groups recruited, H2 carrier frequency varied from 0% to 11.1% (data shown for ethnic groups with H2 present in this cohort in Supplementary Table 2).

**Table 1.** *MAPT* rs1052553 allele/haplotype and genotype distribution in persons with Parkinson’s disease and controls

|  | All participants<br>(n= 1929; 3858 alleles) | Parkinson's disease<br>(n= 907; 1814 alleles) | Control participants<br>(n= 1022; 2044 alleles) | *p* value |
| --- | --- | --- | --- | --- |
| *MAPT* rs1052553 alleles (haplotype) |  |  |  |  |
| A (H1) | 3816 (98.9%) | 1790 (98.7%) | 2026 (99.1%) | 0.19 |
| G (H2)* | 42 (1.1%) | 24 (1.3%) | 18 (0.9%) |  |
| *MAPT* rs1052553 genotypes |  |  |  |  |
| A/A (H1/H1) | 1888 (97.9%) | 884 (97.5%) | 1004 (98.2%) | 0.24 |
| A/G (H1/H2) | 40 (2.1%) | 22 (2.4%) | 18 (1.8%) | 0.31 |
| G/G (H2/H2) | 1 (0.05%) | 1 (0.1%) | 0 (0.0%) | 0.47** |
| *MAPT* rs1052553 genotype category |  |  |  |  |
| H1/H2 (A/G) and H2/H2 (G/G) | 41(2.1%) | 23 (2.5%) | 18 (1.8%) | ref |
| H1/H1 (A/A) only | 1888 (97.9%) | 884 (97.5%) | 1004 (98.2%) | 0.23** |

**Footnote:** Allele frequencies calculated with total number of alleles as denominator (n=3858 overall, 1814 for PD and 1022 for controls). Using the number of participants with one or two of any specified haplotypes (n = total number of participants in category), H2 haplotype frequency = 41/1929 i.e. 2.1% overall, 23/907 (2.5%) in PD and 18/1022 (1.76%) in controls. \*One PD participant homozygous H2 (H2/H2); \*\*p value for Fisher's Exact test. Odds ratio (95% confidence interval) for comparison: 0.68 (0.39 – 1.28). *MAPT* H1/H1 frequency by gender (n vs total): PD (females 251/258, 97.3%; males 633/649, 97.5%) and Controls (females 348/351, 99.2%; males 656/671, 97.8%); p>0.05.

### Association between *MAPT* HI haplotype and PD risk and age of onset

There was no association between H1 haplotype and PD even after adjusting for age at onset of PD and gender (Table 2). The mean age at onset of PD did not differ significantly between individuals with the H1/H1 genotype (60.0 ± 10.6 years) and those with either H1/H2 or H2/H2 (60.0 ± 11.5 years) (p=0.98) as shown in Table 3.

**Table 2.** *MAPT* rs1052553 genotype and risk of Parkinson’s disease

| **Genotype** | **PD**<br>**n = 907 (%)** | **Controls**<br>**N = 1022 (%)** | **Odds ratio (95%CI)** | **\*p-value** |
| --- | --- | --- | --- | --- |
| H1/H2 and H2/H2 | 23 (2.5) | 18 (1.8) | ref |  |
| H1/H1 | 884 (97.5) | 1004 (98.2) | 0.68 (0.39 - 1.28) | 0.23 |

\*Adjusted for gender, age at onset for PD cases and age at study for controls.

**Table 3.**
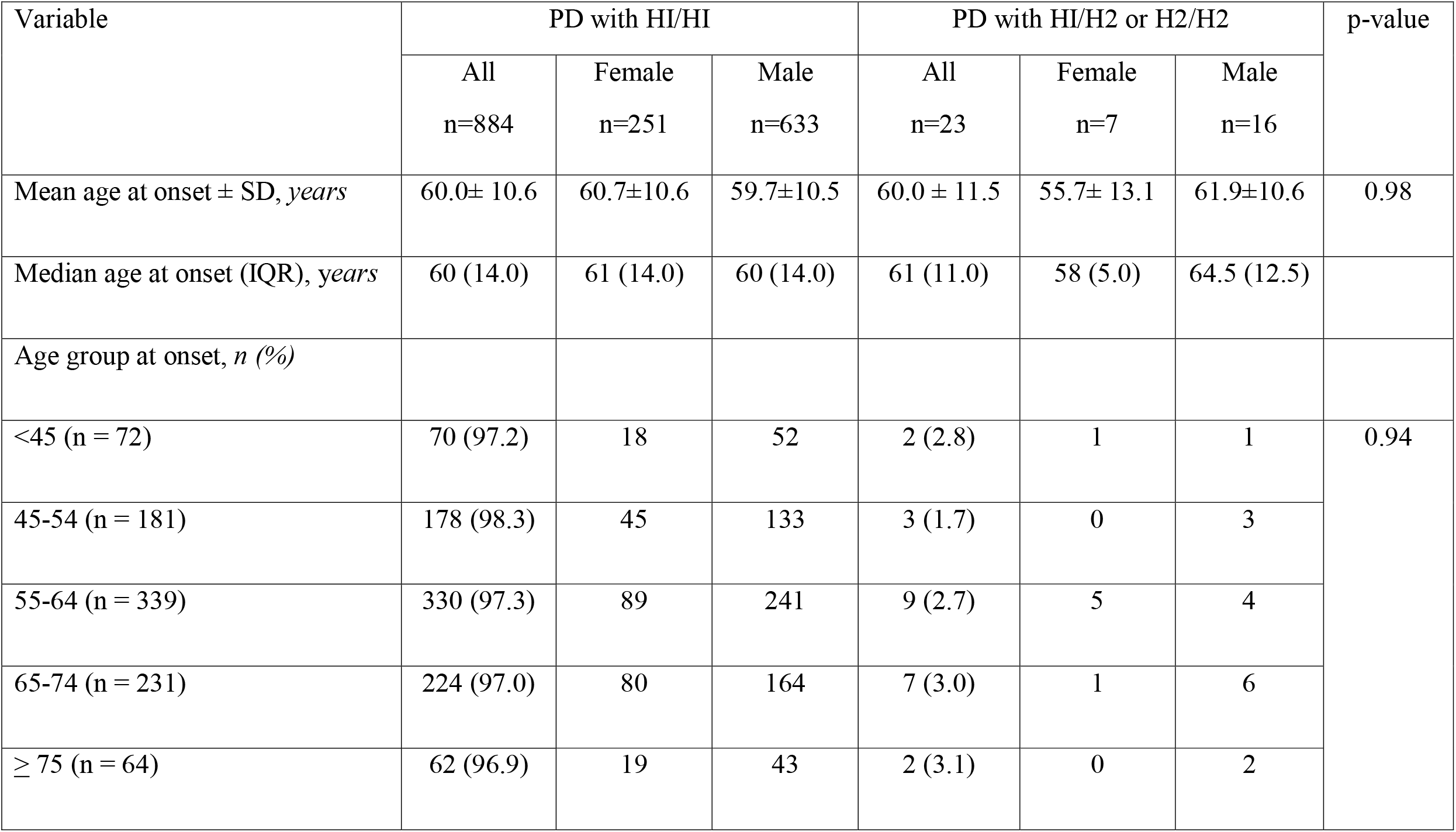
*MAPT* rs1052553 genotype and age at onset in Parkinson’s disease

## Discussion

The likelihood that variations in *MAPT* expression contribute to the pathogenesis of PD nominate it as a target of interest for studies exploring the role of genetic variability in the causation of apparently sporadic PD. Previous studies exploring the *MAPT*-PD connection including those demonstrating an independent and significant effect have been conducted in populations of predominantly Caucasian ancestry, with minimal to no data on individuals of black African ancestry. Our study addresses this gap, and additionally provides important data from a more general perspective on the population genetics of *MAPT* by illustrating the distribution of the 17q21 inversion in individuals of African ancestry from a modestly sized cohort.

*MAPT* H2 haplotype distribution varies widely between sub Saharan Africa and North Africa, Europe and Asia, with the highest levels reported from the ‘Near East’ and Europe. ^3,18^ This difference has been attributed to a genetic drift by founder events affecting the H2D haplotype which possibly occurred in the late Paleolithic or early Neolithic period in the western Near East.^18^ Donnelly *et al* previously described the distribution of the inverted (H2) haplotype using 21 SNPs including the rs1052553 utilized in our study. The study included populations within Africa, consisting of 3 of the Nigerian ethnic groups included in this study (Ibo, Yoruba and Hausa). ^3^ The study concluded that the H2 inverted haplotype occurs at low frequencies in Africa and was completely absent in the Nigerian sample.^3^ Although our findings largely corroborate the low frequency reported in west Africans, we have in fact demonstrated that the H2 haplotype does exist in the Nigerian population, and occurred in 2.1% of our participants. The frequencies for other African populations range between 0.75 to 6.3%, whereas it is reported as occurring in 4% of African Americans.^3, 19^ Donnelly and colleagues had reported that within Africa, the H2 haplotype occurs at the highest frequency in North Africa (Mozabites), at low levels in Northwest Africa, Central Africa, and Eastern Africa, and is absent in West Africa (where Nigeria is located) except in the Mandenka.^3^ Steinberg and colleagues made a similar assertion as, in their study that included Nigerian Yorubas, the H2 inverted haplotype was absent from virtually all Western African individuals except for the Pygmy populations (Bakola, Biaka, and Mbuti).^19^

Regarding the risk of PD conferred by the *MAPT* H1 haplotype, some previous reports indicated an increased risk, and increased transcriptional activity of H1 in comparison to H2 implying a possible role in the development of PD.^4, 9-12, 20, 21^ Increased proportions of H1 resulting in tau mediated alpha-synuclein fibrillisation have been linked to increased PD susceptibility and disease progression.^22^ In contrast to these reports we did not observe any association between H1 and PD risk, a finding similar to some other studies. ^23-25^ Winkler and colleagues have suggested that the role of H1 haplotypes in PD aetiology may be ethnically dependent, based on variable results obtained from their study of two large European groups from Germany and Serbia.^23^ In that study, H1/H1 genotype was significantly associated with PD in only the Serbian subgroup.^23^ Similarly, Davis *et al* did not find an association between *MAPT* variants and PD risk in their predominantly Caucasian North American dataset, but noted an effect on age at onset of PD, suggesting that the influence may be mediated through interactions with other genetic risk variants, including several genes co-located within the H1/H2 haplotype on chromosome 17q21.^24, 26^ A conceivable explanation of the inconsistencies is the observation that distinct subhaplotypes of H1 (such as H1p) (which may be rare or variably distributed in different ethnic populations) may drive the association of *MAPT* with PD (and PD clinical characteristics such as PD dementia) and other neurodegenerative conditions. ^26-28^ Furthermore, relatively small numbers of participants with the H2 haplotype in our population may underlie the inability to corroborate previous reports of a protective effect on risk of PD.^23^

We acknowledge the main limitation of our study in not dissecting the *MAPT* sub-haplotypes and thus limiting the conclusiveness of drivers of the lack of association observed in this study. Whilst our conclusions are credible regarding this lack of association, it is possible that sub-haplotype analysis would have identified the predominant sub-haplotypes in our population and demonstrated any differences with the sub-haplotypes driving PD risk in other populations. Our data represent the first evaluation of the association of *MAPT* and PD in the black African population using a modest sample size comparable to reports from other populations, and re-emphasizes the importance of studying diverse populations.

## Data Availability

All data produced in the present study are available upon reasonable request to the authors

## Acknowledgements

We are very grateful to all our participants and their carers. Funding support was provided by the Michael J Fox Foundation Genetic Diversity in Parkinson’s Disease 2019 (Grant ID:17483) awarded to N.O., M.R., H.H., J.H. and A.S., and the Tertiary Education Trust Fund (TETFund) National Research Fund (NRF) Intervention 2019 (Batch 6) (N.O.).

## Author contributions

Conceptualization and design: N.O., O. Okunoye, O. Ojo, M.R. Data acquisition, analysis or interpretation: All authors. Drafting of manuscript: N.O., O. Okunoye, O. Ojo. Critical revision of manuscript for intellectual content: All authors. Statistical analysis: N.O., O. Okunoye, O. Ojo, S.B-C. Obtained funding: N.O., M.R., H.H., J.H., A.S. Administrative, technical or material support: N.O., O. Ojo, M.R. Supervision: N.O., O. Ojo, M.R. Supervision of genotyping: M.R., D.H. Data management: N.O., O. Okunoye, O. Ojo.

## Competing interests

R.A. is supported by the following grants: US NIH/ NHGRI (U01HG010273) and the UK Royal Society /African Academy of Sciences (FLR/R1/ 191813). M.R. received funding from the University College London Grand challenges Small Grants (Award ID:177813 and the Michael J Fox Foundation Genetic Diversity in Parkinson’s Disease 2019 (Grant ID:17483). H.H. is supported by the Michael J Fox Foundation Genetic Diversity in Parkinson’s Disease (Grant ID: 17483). N.U.O. is supported by the Michael J Fox Foundation Genetic Diversity in Parkinson’s Disease 2019 (Grant ID:17483) and the TETFund National Research Fund (NRF) 2019. S.B.-C., C.B. and A.S. are supported by the Intramural Research Program, National Institute on Aging, National Institutes of Health and US Department of Health and Human Services project ZO1 AG000949 and all these authors declare no non-financial competing interests. The remaining authors declare no competing interests.

## Notes

### Funding Statement

This study was funded by the Michael J Fox Foundation for Parkinson's Research

### Author Declarations

The National Health Research Ethics Committee of Nigeria gave ethical approval for this work.

## References

1. Guo T, Noble W, Hanger DP. Roles of tau protein in health and disease. Acta Neuropathol. 2017;133(5):665–704.

2. Zhang CC, Xing A, Tan MS, Tan L, Yu JT. The Role of MAPT in Neurodegenerative Diseases: Genetics, Mechanisms and Therapy. Mol Neurobiol. 2016;53(7):4893–904.

3. Donnelly MP, Paschou P, Grigorenko E, Gurwitz D, Mehdi SQ, Kajuna SL, et al. The distribution and most recent common ancestor of the 17q21 inversion in humans. Am J Hum Genet. 2010;86(2):161–71.

4. Zhang CC, Zhu JX, Wan Y, Tan L, Wang HF, Yu JT, et al. Meta-analysis of the association between variants in MAPT and neurodegenerative diseases. Oncotarget. 2017;8(27):44994–5007.

5. Zhang H, Cao Y, Ma L, Wei Y, Li H. Possible Mechanisms of Tau Spread and Toxicity in Alzheimer’s Disease. Front Cell Dev Biol. 2021;9: 707268. doi: 10.3389/fcell.2021.707268.

6. Silva MC, Haggarty SJ. Tauopathies: Deciphering Disease Mechanisms to Develop Effective Therapies. Int J Mol Sci. 2020;21(23):8948. doi: 10.3390/ijms21238948.

7. Wade-Martins R. The MAPT locus—a genetic paradigm in disease susceptibility. Nature Reviews Neurology. 2012;8(9):477–8.

8. International Parkinson Disease Genomics Consortium; Nalls MA, Plagnol V, Hernandez DG, Sharma M, Sheerin UM, et al. Imputation of sequence variants for identification of genetic risks for Parkinson’s disease: a meta-analysis of genomewide association studies. Lancet. 2011;377(9766):641–9.

9. Ezquerra M, Pastor P, Gaig C, Vidal-Taboada JM, Cruchaga C, Muñoz E, et al. Different MAPT haplotypes are associated with Parkinson’s disease and progressive supranuclear palsy. Neurobiology of Aging. 2011;32(3):547.e11-e16.

10. Mata IF, Yearout D, Alvarez V, Coto E, de Mena L, Ribacoba R, et al. Replication of MAPT and SNCA, but not PARK16-18, as susceptibility genes for Parkinson’s disease. Mov Disord. 2011;26(5):819–23.

11. Elbaz A, Ross OA, Ioannidis JP, Soto-Ortolaza AI, Moisan F, Aasly J, et al. Independent and joint effects of the MAPT and SNCA genes in Parkinson disease. Ann Neurol. 2011;69(5):778–92.

12. Vandrovcova J, Pittman AM, Malzer E, Abou-Sleiman PM, Lees AJ, Wood NW, et al. Association of MAPT haplotype-tagging SNPs with sporadic Parkinson’s disease. Neurobiol Aging. 2009;30(9):1477–82.

13. Huang Y, Wang G, Rowe D, Wang Y, Kwok JB, Xiao Q, et al. SNCA Gene, but not MAPT, influences onset age of Parkinson’s Disease in Chinese and Australians. Biomed Res Int. 2015;2015:135674. doi: 10.1155/2015/135674.

14. Ojo OO, Abubakar SA, Iwuozo EU, Nwazor EO, Ekenze OS, Farombi TH, et al. The Nigeria Parkinson Disease Registry: Process, Profile, and Prospects of a Collaborative Project. Mov Disord. 2020;35(8):1315–1322.

15. Rizig M, Okubadejo N, Salama M, Thomas O, Akpalu A, Gouider R; IPDGC Africa. The International Parkinson Disease Genomics Consortium Africa. Lancet Neurol. 2021;20(5):335.

16. Hughes AJ, Daniel SE, Kilford L, Lees AJ. Accuracy of clinical diagnosis of idiopathic Parkinson’s disease: a clinico-pathological study of 100 cases. J Neurol Neurosurg Psychiatry. 1992;55(3):181–4.

17. Valenca GT, Srivastava GP, Oliveira-Filho J, White CC, Yu L, Schneider JA, et al. The role of MAPT haplotype H2 and isoform 1N/4R in parkinsonism of older adults. PLoS One. 2016;11(7):e0157452.

18. Espinosa I, Alfonso-Sánchez MA, Gómez-Pérez L, Peña JA. Neolithic expansion and the 17q21.31 inversion in Iberia: an evolutionary approach to H2 haplotype distribution in the Near East and Europe. Mol Genet Genomics. 2023;298(1):153–160.

19. Steinberg KM, Antonacci F, Sudmant PH, Kidd JM, Campbell CD, Vives L, et al. Structural diversity and African origin of the 17q21.31 inversion polymorphism. Nat Genet. 2012;44(8):872–80.

20. Li J, Ruskey JA, Arnulf I, Dauvilliers Y, Hu MTM, Högl B, et al. Full sequencing and haplotype analysis of MAPT in Parkinson’s disease and rapid eye movement sleep behavior disorder. Mov Disord. 2018;33(6):1016–1020.

21. Kwok JB, Teber ET, Loy C, Hallupp M, Nicholson G, Mellick GD, et al. Tau haplotypes regulate transcription and are associated with Parkinson’s disease. Ann Neurol. 2004;55(3):329–34.

22. Giasson BI, Forman MS, Higuchi M, Golbe LI, Graves CL, Kotzbauer PT, et al. Initiation and synergistic fibrillization of tau and alpha-synuclein. Science. 2003;300(5619):636–40.

23. Winkler S, König IR, Lohmann-Hedrich K, Vieregge P, Kostic V, Klein C. Role of ethnicity on the association of MAPT H1 haplotypes and subhaplotypes in Parkinson’s disease. Eur J Hum Genet. 2007;15(11):1163–8.

24. Davis AA, Andruska KM, Benitez BA, Racette BA, Perlmutter JS, Cruchaga C. Variants in GBA, SNCA, and MAPT influence Parkinson disease risk, age at onset, and progression. Neurobiol Aging. 2016;37:209.e1-209.e7.

25. Miranda-Morales EG, Sandoval-Carrillo A, Castellanos-Juárez FX, Méndez-Hernández EM, La Llave-León O, Quiñones-Canales G, et al. H1/H2 MAPT haplotype and Parkinson’s disease in Mexican mestizo population. Neurosci Lett. 2019;690:210–213.

26. Bowles KR, Pugh DA, Liu Y, Patel T, Renton AE, Bandres-Ciga S; International Parkinson’s Disease Genomics Consortium (IPDGC), et al. 17q21.31 sub-haplotypes underlying H1-associated risk for Parkinson’s disease are associated with LRRC37A/2 expression in astrocytes. Mol Neurodegener. 2022;17(1):48. doi: 10.1186/s13024-022-00551-x.

27. Setó-Salvia N, Clarimón J, Pagonabarraga J, Pascual-Sedano B, Campolongo A, Combarros O, et al. Dementia risk in Parkinson disease: disentangling the role of MAPT haplotypes. Arch Neurol. 2011 Mar;68(3):359–64.

28. Heckman MG, Kasanuki K, Brennan RR, Labbé C, Vargas ER, Soto AI, et al. Association of MAPT H1 subhaplotypes with neuropathology of lewy body disease. Mov Disord. 2019;34(9):1325–1332.

