## Supplementary Tables for "*MAPT* allele and haplotype frequencies in Nigerian Africans: population distribution and association with Parkinson’s disease risk and age at onset"

**Supplementary Materials**

**Supplementary Table 1. Baseline characteristics of individuals with PD and controls**

| **Variables** | **All participants**  **n = 1929** | **PD**  **n=907** | **Controls**  **n=1,022** | **p-value** |
| --- | --- | --- | --- | --- |
| Female | 609 (31.6) | 258 (28.4) | 351 (34.3) | 0.005 |
| Male | 1320 (68.4) | 649 (71.6) | 671 (65.7) |  |
| Mean age at study (±SD), *years* | 63.8 ±9.8 | 64.1 ± 10.2 | 63.4 ± 9.3 | 0.10 |
| Median age at study (IQR), *years* | 64.0 (12) | 65.0 (13) | 63.0 (12) |  |
| Mean age at study (±SD) (females), *years* | 63.6 ±9.5 | 64.9 ±9.8 | 62.6 ±9.1 |  |
| Mean age at study (±SD) (males), *years* | 63.8±9.9 | 63.9±10.4 | 63.8±9.4 |  |
| Mean age at onset (±SD), *years* | N/A | 60.0 ±10.6 | N/A |  |
| Median age at onset (IQR), *years* | N/A | 60.0 (14.0) | N/A |  |
| Mean age at onset (±SD) (females), *years* | N/A | 60.6 ±10.7 | N/A | 0.25 |
| Mean age at study (±SD) (males), *years* | N/A | 59.7 ±10.5 | N/A |  |
| Duration of disease (mean ±SD), years | N/A | 4.2 ±4.1 | N/A |  |
| Median duration of disease (IQR), years | N/A | 3.0 (3.0) | N/A |  |
| Duration of disease (mean ±SD) (females), years | N/A | 4.3 ±4.5 | N/A | 0.73 |
| Duration of disease (mean ±SD) (males), years | N/A | 4.2 ±3.9 | N/A |  |

**Supplementary Table 2. Distribution of H2 haplotype by Nigerian ethnic group in the entire study cohort**

| Self-declared ethnic group | Number of participants | H2 carriers  n (%) |
| --- | --- | --- |
| Yoruba | 684 | 8 (1.2) |
| Igbo | 381 | 21(5.5) |
| Hausa | 247 | 2 (0.8) |
| Other (self-named ethnic group) | 202 | 4 (1.98) |
| Ibibio | 55 | 1 (1.8) |
| Idoma | 43 | 1 (2.3) |
| Efik | 36 | 1 (2.8) |
| Fulani | 25 | 2 (8.0) |
| Anang | 9 | 1 (11.1) |
| Tiv | 76 | 0 (0) |
| Ishan/Esan | 30 | 0 (0) |
| Urhobo | 29 | 0 (0) |
| Bini | 19 | 0 (0) |
| Igalla | 17 | 0 (0) |
| Birom | 15 | 0 (0) |
| Kanuri | 13 | 0 (0) |
| Isoko | 9 | 0 (0) |
| Itsekiri | 9 | 0 (0) |
| Igbirra | 7 | 0 (0) |
| Nupe | 7 | 0 (0) |
| Ukwani | 7 | 0 (0) |
| Etsako | 4 | 0 (0) |
| Jukun | 2 | 0 (0) |
| Shuwa | 2 | 0 (0) |
| Iyalla | 1 | 0 (0) |
| Total | 1929 | 41 (2.1%) |

Footnote: Ethnic groups arranged by sample size within cohort (highest to lowest) and frequency of H2 haplotype. Number of participants includes entire cohort of persons with PD and controls.
